# TUMOUR NECROSIS FACTOR ALPHA GENE PROMOTER POLYMORPHISM IN PATIENTS WITH SCHIZOPHRENIA IN A KENYAN POPULATION

**DOI:** 10.1101/2025.05.18.25327849

**Authors:** Mowlem Pierre, Isaac Ndede, Lukoye Atwoli

**Author notes:** **Correspondence:** Mowlem Pierre, Department of Pathology, Moi University School of Medicine, Eldoret, Kenya;.

## Abstract

**Aim:** Tumour necrosis factor alpha (TNF-α) promoter single nucleotide polymorphisms (SNPs) have been reported to play pathological roles in schizophrenia with contradicting findings in several studies. The status of TNF-α SNPs remains unknown in our population. Therefore, this study sought to determine the association of TNF-α gene promoter single nucleotide polymorphisms -308A/G (rs 1800629) and -1031C/T (rs 1799964) in patients with schizophrenia at MTRH, Kenya.

**Methods:** A case-control study at MTRH included 82 schizophrenia patients and 82 age and sex-matched healthy controls. Participants’ demographic data were collected using an interviewer-administered questionnaire, and DNA was extracted from saliva samples for TNF-α SNP genotyping using the Allelic Discrimination Assay®.

**Results:** There were no statistically significant differences in genotype distribution and allele frequencies between cases and controls for -308A/G and -1031C/T polymorphisms (p = 0.261) and (p = 0.678) respectively. The genotype associations with schizophrenia were: at - 308(rs1800629), A/G had OR 1.67, 95% CI (0.88-3.16) p=0.114. At -1031(rs1799964), genotype C/C and C/T, had OR 0.66, 95% CI (0.22-1.97) p=0.455 and OR 0.75, 95% CI (0.48-1.18) p=0.255 while a combination of C/T-C/C, had OR 0.743, 95% CI (0.40-1.38) p=0.346.

**Conclusion:** These results demonstrate that SNPs in the TNF-α promoter region at the studied loci are unrelated to schizophrenia. These SNPs may not serve as useful biomarkers for diagnosis or targeted therapy. Further analysis of these SNPs focusing on specific schizophrenia symptoms, rather than the entire syndrome, may be needed.

## Introduction

Schizophrenia is a complex mental disorder that is characterized by abnormalities of thought, emotion, and behaviour [1], with a global prevalence rate of 1.4 - 4.6 per thousand population and is associated with significant healthcare cost and functional decline [2]. Not only is schizophrenia the most common psychosis but it also tends to involve abnormalities in all of the symptom clusters such as positive, negative, mood symptoms and cognitive deficits that tend to reliably develop during late adolescence and early adulthood about 15 - 35 years [3]. Nevertheless, schizophrenia is recognized as a systemic syndrome, involving not only the nervous system but also the immune system, as well as originating from a combination of environmental and genetic factors [4]. The exact cause of schizophrenia remains unknown, with its complex pathophysiology influenced by interactions between the brain and other organ systems. Various hypotheses, including genetic, neurodevelopmental, neurotransmitter, neuroimmunological factors, and abnormalities in the immune system and central nervous system particularly involving cytokines have been proposed to explain its aetiology and pathogenesis [1]. No single factor can be identified as the sole cause of schizophrenia, as research has primarily focused on neurotransmitter systems, especially dopamine [5]. Inflammatory cytokines play a role by affecting the synthesis and uptake of monoamine neurotransmitters, including dopamine, serotonin, and norepinephrine, as well as influencing their release [6]. The exact aetiology and underlying biology of schizophrenia are still poorly understood, with diagnosis based primarily on clinical symptoms rather than pathophysiology. Both genetic factors and environmental influences, such as trauma and stress, as well as disturbances in immune processes involving inflammatory cytokines like IL-6, IL-1, IL-8, and TNF-α, have been linked to the disorder [7]. TNF-α plays a complex role in schizophrenia, influenced by genetic polymorphisms in the genes that regulate its production. Certain variations or polymorphisms in the TNF-α promoter region can lead to increased TNF-α levels [8]. Several single nucleotide polymorphisms (SNPs) in the TNF locus can affect transcription and protein production of TNF-α, influencing disease outcomes [9]. Notably, polymorphisms in the promoter region, such as those at positions -308 and -1031, may alter TNF-α levels in both the peripheral and central nervous systems, contributing to imbalances in pro-inflammatory cytokines during disease processes [10]. TNF-α acts by binding to two receptors on target cells: TNFR 1 and TNFR 2. This binding activates pathways that promote cell survival and trigger inflammatory responses, including the activation of NF-κB and MAPK. These processes enhance phagocyte activity to clear infections and debris and facilitate the movement of immune cells, such as neutrophils and macrophages, to sites of tissue damage and infection [8]. The TNF genes are found in the Class III region of the human major histocompatibility complex (MHC) on chromosome 6p21.1 - 6p21.3, and they exhibit a high number of polymorphisms compared to other cytokine genes. Some of these polymorphisms create extended haplotypes with HLA class I and II alleles, making TNF gene and promoter polymorphisms relevant in various diseases, particularly those associated with HLA [11]. The exact relationship between specific SNPs of the TNF-α inflammatory cytokine and schizophrenia remains unclear in our population.

Research on TNF-α gene promoter polymorphisms in schizophrenia is notably limited in African populations, particularly in Kenya. To our knowledge to date, no study has assessed the -308 A/G and -1031 C/T SNPs in schizophrenia in the African populations in particular Kenya. This study aims to explore the relationship between TNF-α gene promoter polymorphisms and schizophrenia among patients at Moi Teaching and Referral Hospital.

## Materials and Methods

### Study design

This was a case-control study nested in the larger Neuropsychiatric Genetics of African Population-Psychosis (NeuroGAP-P) study in African populations [12], conducted at the Moi Teaching and Referral Hospital (MTRH) Psychiatric Clinic. The study focused on cases who where patients seeking or referred for treatment at the outpatient Psychiatric clinic at MTRH, with a clinical diagnosis of schizophrenia based on a Structured Clinical Interview for DSM-V by experienced psychiatrists and Controls comprised of healthy individuals drawn from the MTRH facility as caretakers accompanying patients, students, and staff or for other non-psychiatric purposes persons with no signs of mental and infectious illnesses as determined by clinical evaluations and self-reports. The study starts from August 2021 until April 2022. The research complied with ethical standards with approvals from the Institutional Research and Ethics Committee of Moi University/Moi Teaching and Referral Hospital, Eldoret, Kenya (approval number MU/MTRH-FAN:0003881).

### Sample collection procedure

Participants were invited to participate in the study if they met the inclusion criteria. Inclusion of cases was based on clinical diagnosis of Schizophrenia confirmed by a Psychiatrist, either male or female at least 18 years of age with willingness and ability to participate. Exclusion criteria for patients were those exhibiting intrusive levels of psychiatric symptoms, under the age of 18 years, and having acute levels of alcohol or substance use as demonstrated by being a current patient or under acute medical care of one of these conditions and persons not fluent in one or all of the languages the consent form has been translated into and not being able to consent to the study.

Inclusion of controls consisted of healthy individuals of at least 18 years of age either male or female with no signs of mental illness or obvious signs of infectious diseases as determined by clinical evaluation and self-reports. Exclusion of controls comprised individuals with psychotic disorder diagnosis, acute infections, current or history of psychotic symptoms or are on psychiatric medication. Those having acute levels of alcohol or substance use as demonstrated by being a current inpatient or under acute medical care for one of these conditions and not fluent in one or all of the languages the consent form has been translated into and not being able to consent to the study.

Before enrollment, the participants who met the inclusion criteria and agreed to participate (164 participants) in the study, were provided with a detailed explanation of the study’s purpose and procedure and enrollment into the study followed. Capacity to consent was assessed thereafter, with a review of informed consent and signing of a consent form. Thereafter participant’s demographics and details were captured in a questionnaire after which saliva was collected by spitting in the Oragene Saliva collection kit and stored at room temperature (15-25 ºC) for further examination of genotyping.

### Procedures for genotyping

Genomic DNA was extracted from the saliva of both patients and controls according to the standard procedure of the Ethanol precipitation protocol and prepIT®•L2P reagent (Genotek Inc. Ottawa, ON, Canada). The quality of the DNA extract measurements was checked spectrophotometrically by a NanoDrop™ OneC spectrophotometer (Thermo Fisher Scientific, USA). Genomic DNA extracted from saliva was genotyped for two single nucleotide polymorphisms (SNPs) in the promoter region of the TNF-α gene at nucleotide position -308 A/G (rs 1800629) and -1031 C/T (rs 1799964) using Allelic discrimination assay protocol. Two fluorescent dyes, a reporter and a quencher are attached to the probes with the TaqMan PCR Reagent Kit. When both dyes are attached to the probe, reporter dye emission is quenched. Taq DNA polymerase cleaves the reporter dye from the probe through its 5`-3` exonuclease during each extension cycle. Once separated from the quencher, the reporter dye emits its characteristic fluorescence which can then be measured by the Quantstudio 5 instrument (Applied Biosystems, USA). The amount of fluorescence measured is proportional to the amount of PCR product made. Primers and probes for detection of the SNPs are (−308 A/G) 5′-GGACCCTGGAGGCTGAAC - 3′ (forward primer), 5′ - CCAAAAGAAATGGAGGCAATAGGTT - 3′ (reverse primer), 5′ - VIC - CCCGTCCCCATGCC - NFQ - 3′ (probe 1) and 5′ - FAM - CCCGTCCTCATGCC - NFQ – 3′ (probe 2). For (−1031 C/T) 5′ - GTGAGGCCGCCAGACT – 3′ (forward primer), 5′ - GCCCCTCCAGACCCTGA - 3′ (reverse primer), 5′ - VIC – CTTTTCCTTCGTCTTCTC – NFQ -3′ (probe 1) and 5′ - FAM – TTTTCCTTCATCTTCTC – NFQ - 3′ (probe 2). Thermal cycling conditions for polymerase chain reaction were: pre-read stage 50°C for 2 minutes, hold stage 95°C for 10 min, PCR stage 95°C for 15 seconds and 60°C for 1 min for 40 cycles, and post-read stage 60°C for 30 seconds.

### Data analysis

The data was first cleaned and entered into a Microsoft Excel spreadsheet. The data was then imported and analysed using R-studio version 4.2.2. The demographic data, characteristics of the study participants and laboratory results were described using means, standard deviations and percentages. Kruskal-Wallis and Fisher’s exact test was applied to test the statistical significance of differences between genotypes and allele frequencies in the cases and control groups and conformation to Hardy-Weinberg equilibrium (HWE). Logistic regression estimated by the odds ratio assessed the associations between schizophrenia with particular genotypes at different SNPs -308 A/G (rs 1800629) and -1031 C/T (rs 1799964). For all tests, a p-value < 0.005 was considered statistically significant.

## Results

Genotype distribution for case and control groups for all markers conformed to Hardy-Weinberg equilibrium. The genotype distribution of the TNF-α gene -308 A/G (A/G) heterozygous genotype was higher among the cases at 17.1% (wild type) than the controls at 11% (p = 0.369). The homozygous G/G genotype was higher in controls at 89% compared to the cases at 82.9%. The A/A homozygous genotype was neither identified in the cases nor the control groups. Genotypes on the TNF-α gene promoter at -1031, the T/T homozygous genotypes were higher (59.8%, wild type) among the cases compared to the controls at 52.4%. The controls had higher C/T heterozygous genotypes at 42.7% compared to the cases at 36.6% and higher C/C homozygous genotypes at 4.9% compared to the cases at 3.7%. There was no statistically significant difference (p = 0.678) in the distribution of genotypes between the cases and the controls (Table 1).

**Table 1:**
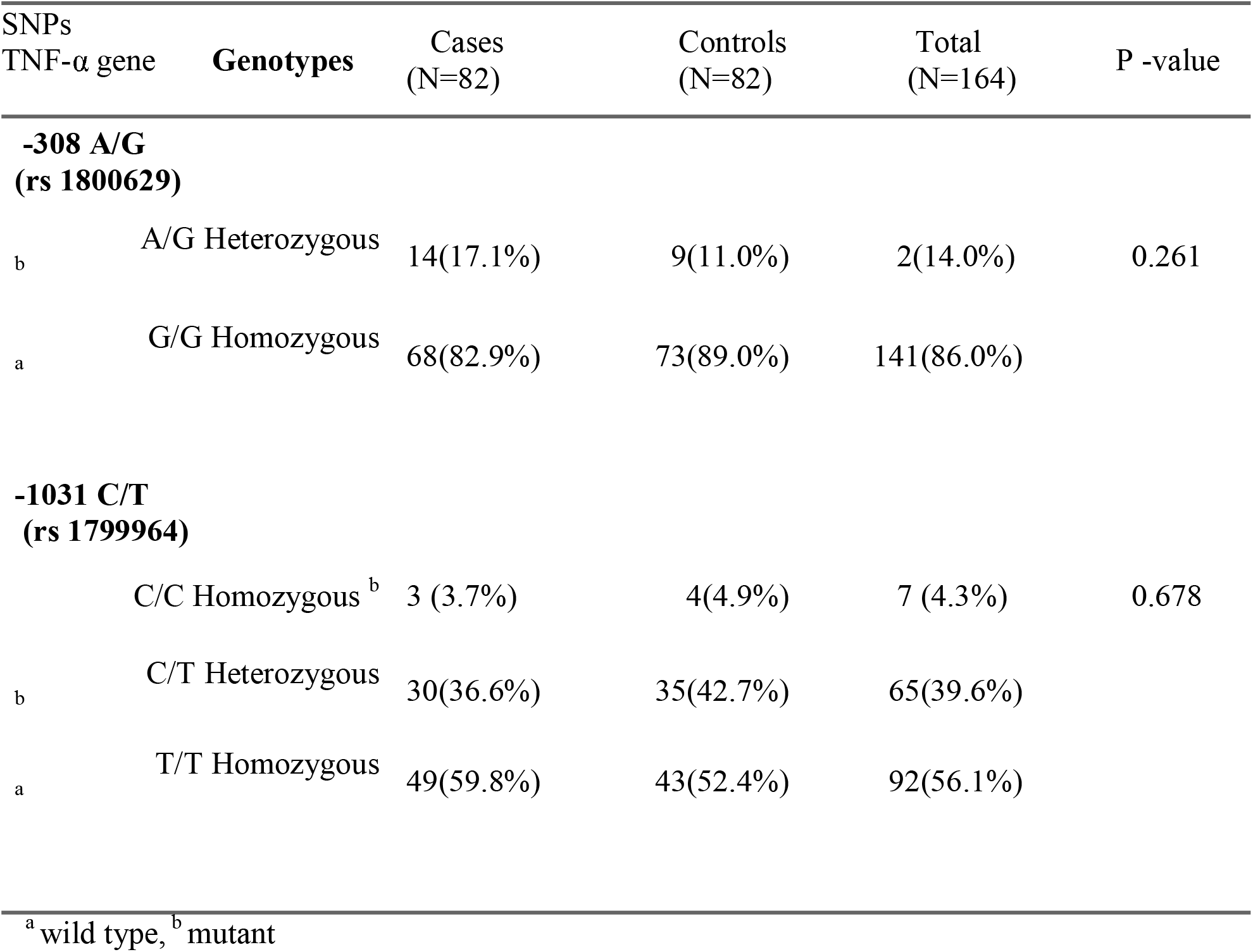
Genotype distribution of the TNF-α gene promoter -308 A/G and -1031 C/T among cases and controls.

| SNPs |  | Cases | Controls | Total | P -value |
| --- | --- | --- | --- | --- | --- |
| TNF-α gene | Genotypes | (N=82) | (N=82) | (N=164) | |
| **-308 A/G** |  |  |  |  |  |
| **(rs 1800629)** |  |  |  |  |  |
| b | A/G Heterozygous | 14(17.1%) | 9(11.0%) | 2(14.0%) | 0.261 |
| a | G/G Homozygous | 68(82.9%) | 73(89.0%) | 141(86.0%) |  |
| **-1031 C/T** |  |  |  |  |  |
| **(rs 1799964)** |  |  |  |  |  |
|  | C/C Homozygous<sup>b</sup> | 3 (3.7%) | 4(4.9%) | 7 (4.3%) | 0.678 |
| b | C/T Heterozygous | 30(36.6%) | 35(42.7%) | 65(39.6%) |  |
| a | T/T Homozygous | 49(59.8%) | 43(52.4%) | 92(56.1%) |  |

<sup>a</sup> wild type, <sup>b</sup> mutant

The Adenine (A) allele frequency of the TNF-α gene promoter -308 A/G, was slightly higher in cases at 8.5% compared to the controls at 5.5%, while the allele guanine (G) was marginally higher in the controls at 94.5% compared to the cases at 91.5% (p = 0.278). At TNF-α gene - 1031 C/T, the allele T was higher in cases at 78% compared to the controls at 73.8%, while the frequency of the allele C was higher in controls at 26.2% compared to the cases at 22% (p = 0.366) (Table 2).

**Table 2:** Allele frequencies of the TNF-α gene promoter -308 A/G and -1031 C/T among cases and controls.

| SNPs<br>TNF-αgene<br>Allele | Cases<br>(N=164) | Controls<br>(N=164) | Total<br>(N=328) | P-value |
| --- | --- | --- | --- | --- |
| **-308 A/G**<br>**(rs 1800629)** |  |  |  |  |
| A | 14 (8.54%) | 9 (5.49%) | 23 (7.01%) | 0.278 |
| G | 150(91.46%) | 155 (94.51%) | 305(92.99%) |  |
| **-1031 C/T**<br>**(rs 1799964)** |  |  |  |  |
| C | 36(21.95%) | 43 (26.22%) | 79<br>(24.06%) | 0.366 |
| T | 128(78.05) | 121 (73.78%) | 249(75.94) |  |

Multiple logistic regression assessed the association of SNP genotype at -308A/G and -1031C/T of the TNF-α gene promoter with schizophrenia. At SNP -308 A/G of the TNF-α gene, G/G homozygous were used as the references. The A/G heterozygous genotype had (OR 1.67, 95% [CI 0.88 - 3.16] p = 0.114), showing no statistically significant association with schizophrenia.

At SNP -1031 C/T of the TNF-α gene, the T/T homozygous genotype was used as the reference to evaluate heterozygous C/T and homozygous C/C genotypes. The T/T homozygous genotype versus C/T heterozygous genotype had (OR 0.75, 95% [CI 0.48 - 1.18] p = 0.215) and the T/T homozygous genotype versus C/C homozygous had (OR 0.66, 95% [CI 0.22 - 1.97] p = 0.455). These genotypes were not statistically significantly associated with schizophrenia. The T/T homozygous genotype was used as the reference, to evaluate the combination of homozygous C/C genotype and C/T homozygous genotypes to determine the association for schizophrenia. The combined C/C and C/T genotypes had (OR 0.743, 95% [CI 0.40 - 1.38] p = 0.346). These genotypes were not statistically significantly associated with schizophrenia (Table 3).

**Table 3:**
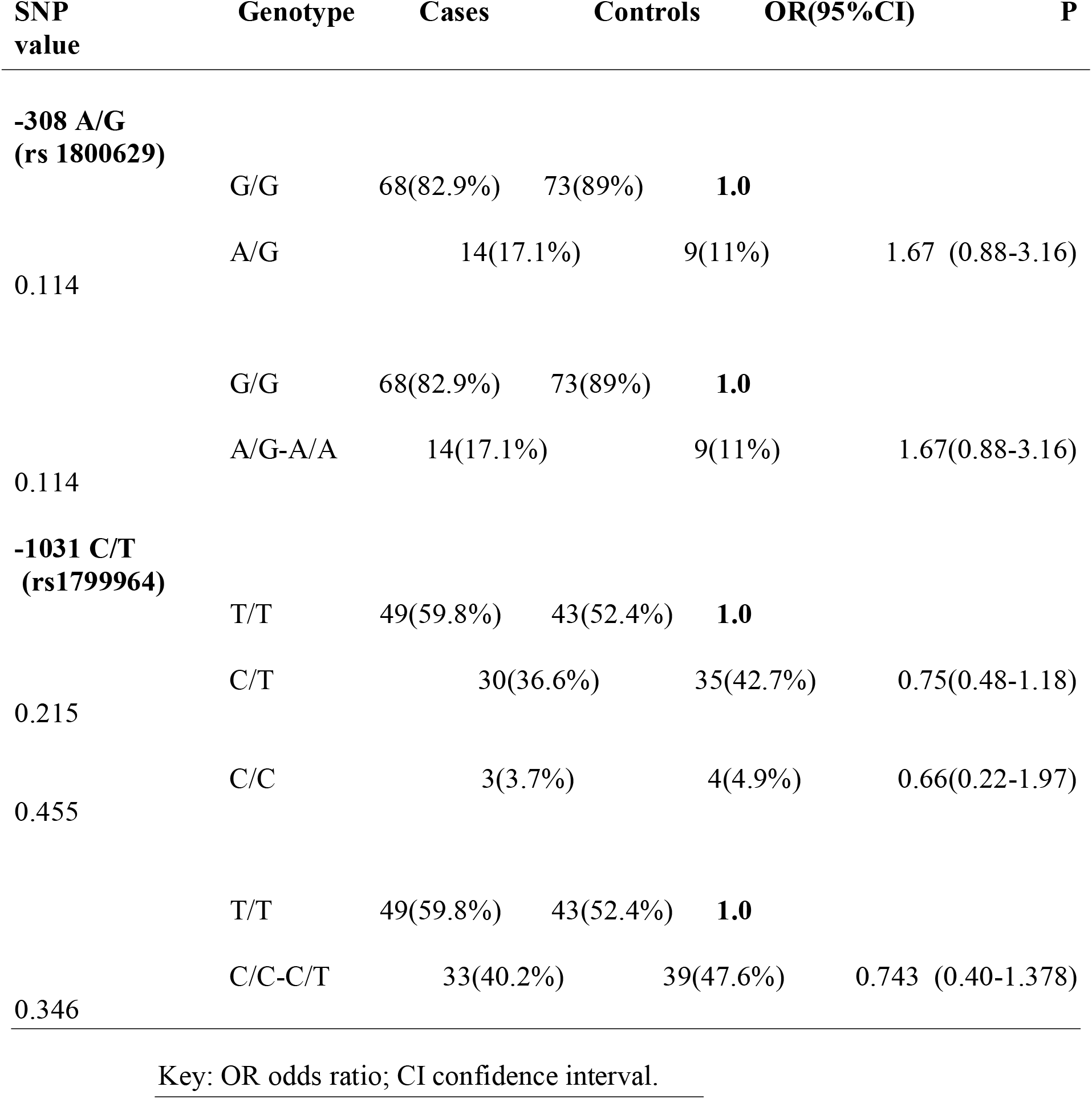
Associations between -308A/G and -1031C/T SNPs of TNF-α gene and schizophrenia under different genotypes.

## Discussion

The study found no association between the TNF-α gene promoter polymorphisms (−308 A/G and -1031 C/T) and schizophrenia in our Kenyan studied population. The genotype distribution at -308 A/G (rs1800629) the heterozygous A/G genotype in the case group had a higher distribution than the control group and homozygous A/A was not detected in either the cases or controls. This was similar in a Chinese population [13] where among the case group, the heterozygous A/G genotypes were high in distribution compared to the control group and the A/A genotype was not detected in the population in either the cases or the control groups. In contrast to a Polish population [14] among the case group, the heterozygous A/G genotype occurred less and the homozygous A/A genotype had a higher distribution than the control group, whereas in the control group, the G/G genotype had a low distribution to the case group. In SNP -1031C/T (rs 1799964) of the TNF-α gene, the cases had a higher distribution of homozygous T/T genotype. These findings were similar to an American Caucasian population [15] where cases had a higher homozygous T/T genotype distribution. Contrary findings were observed in a Chinese population [13] wherein cases had lower heterozygous C/T genotypes. The differences in genotype distribution may be a result of mutation which affects the gene expression and influences the TNF-α expression. These findings may suggest that race differences in a population may account for the variations in the etiological significance of these SNPs in patients with schizophrenia.

On the frequencies of the polymorphism on SNP -308A/G (rs 1800629), the allele A was higher in the cases compared to the controls. These findings were similar to a Chinese population [13] where in the case group the A allele was higher in frequency compared to the controls. However, contradicting findings were observed in a Japanese population [16] wherein the control group, the allele A had a higher frequency compared to the case group. On the frequencies of the polymorphism on SNP -1031C/T (rs1799964), allele T occurred most in the cases compared to the control group. These findings were similar to a case-control study in a Chinese Han population [17] where cases had a high allele T frequency compared to the control group. Contrary findings were observed in a Singaporean population [18] where the allele T had a lower frequency in the case group than in the control group. These differences may be due to the presence of rare variants of alleles at this TNF-α gene. In contrast, the variations in allele frequencies among the case and control groups may have been a result of genetic drift, where the variants in allele frequencies may be an indication of alteration of transcription activities at the promoter, which affects the expression of genes at the transcription and translation levels. We found no significant association between -308A/G and -1031C/T with schizophrenia. These results are in line with the negative findings in the Taiwanese sample [19] and a meta-analysis respectively [20]. However, associations were observed in both -308A/G and -1031C/T in a Caucasian Polish population [1]. The difference in association of promoter SNPs of the TNF-α gene with schizophrenia may arise due to how other potential confounding factors were addressed. In this study, only the direct association between SNPs and schizophrenia was analyzed, while other authors have stratified their cohorts to analyze the relationship between SNPs and specific psychotic symptoms. Since schizophrenia is a complex collection of a variety of symptoms, it would be very difficult to find an association between the broad collection of symptoms and certain SNPs associated with immune markers in a specific location. However, there is the possibility that these polymorphisms could be associated with schizophrenia in non-African populations.

We conclude that the promoter SNP TNF-α genes -308A/G and -1031C/T occur but appear to be not associated with schizophrenia in the studied Kenyan population. However other studies indicate that polymorphisms of TNF-α gene might be associated with schizophrenia. Further studies of single nucleotide polymorphisms at loci other than -308A/G (rs 1800629) and -1031 C/T (rs 1799964) may be needed to draw a more comprehensive association with more specific symptoms of schizophrenia rather than the whole syndrome.

## Data Availability

All data produced in the present study are available upon reasonable request to the authors.

## Abbreviations

DSM-V: Diagnostic Statistical Manual -Version 5
HLA: Human Leukocyte Antigen
IL: Interleukin
MAPK: Mitogen-activated protein kinase
PCR: Polymerase Chain Reaction
NFQ: Non-Fluorescent Quencher
TNFR: Tumor Necrosis Factor Receptor

## Declaration

All data produced in the present study are available upon reasonable request to the authors.

## Author contribution

MP: Conceptualization, performed experiments, writing-original draft, IN: Conceptualization, writing - review &editing, Supervision, LA: Resources, Writing-review and editing, Supervision. All authors read and approved the submitted version.

## Ethical Approval

Ethical approval for the research was granted by the Moi University/ Moi Teaching and Referral Hospital Institutional Research and Ethics Committee (approval number MU/MTRH-FAN: 0003881). Additionally, permission to conduct the study was granted by the CEO of Moi Teaching and Referral Hospital.

## Consent to Participate

Written informed consent to participate in the study was obtained from all the study participants.

## Consent to Publication

Not applicable.

## Availability of Data material

The raw data summarized in this manuscript will be made available by the corresponding author upon request.

